# Occupational exposures associated with being a COVID-19 case; evidence from three case-controls studies

**DOI:** 10.1101/2020.12.21.20248161

**Authors:** Iina Hiironen, Maria Saavedra-Campos, Jennifer Panitz, Thomas Ma, Olisaeloka Nsonwu, Andre Charlett, Gareth Hughes, Isabel Oliver

## Abstract

**Background:** The evidence on risk factors for transmission of SARS-CoV-2 in community settings is sparse, yet this information is key to inform public health action. We investigated factors associated with being a COVID-19 case using data collected through contact tracing.

**Methods:** We conducted three retrospective, frequency-matched case-control studies between August 2020 and October 2020 using case data from the NHS Test and Trace programme. Controls were obtained through Market Research Panels. Multivariable analyses provided adjusted odds ratios (aORs) for multiple community exposure settings. We analysed the results in meta-analyses using random effects models to obtain pooled odds ratios (pORs).

**Results:** Across all study periods, there was strong statistical evidence that working in healthcare (pOR 2.87, aOR range 2.72-3.08), social care (pOR 4.15, aOR range 2.46-5.41) or hospitality (pOR 2.36, aOR range 2.01-2.63) were associated with increased odds of being a COVID-19 case. There was also evidence that working in warehouse setting was associated with increased odds (pOR 3.86, aOR range 1.06-14.19), with a substantial increase in odds observed over the study periods. A similar pattern was also observed in education and construction.

**Conclusions:** The studies indicate that some workplace settings are associated with increased odds of being a case. However, it is not possible to determine how much of the transmission of SARS-CoV-2 took place within the workplace, and how much was associated with social, household or transport exposures.

## Background

Non-pharmaceutical interventions have played a critical role in the COVID-19 response, and are likely to remain core interventions for the foreseeable future despite the promising advances in vaccination programmes. Governments and Public Health authorities rely on the evidence on factors associated with transmission of SARS-CoV-2 to inform control programmes including restrictions to activities. Transmission is determined by contact patterns, environmental and socioeconomic factors (Cevik et al., 2020). It can occur in most settings, but some settings are likely to facilitate or amplify the risk of transmission due to a combination of behavioural and environmental factors (Meyerowitz et al., 2020). Situations in which there is close-proximity contact, contact that is sustained over a prolonged period of time or multiple contacts in a confined poorly ventilated space pose the greatest risk for SARS-CoV-2 transmission (Cevik et al., 2020). There is growing evidence that the risk of COVID-19 is highest in household settings, and that living in an overcrowded or multiple occupancy household increases the risk of becoming a COVID-19 case (Lee et al., 2020).

There is also an increasing amount of evidence on the risk of COVID-19 among healthcare workers (Nguyen et al., 2020; ECDC 2020; Leclerc et al., 2020). Certain community settings have also been associated with transmission. Hospitality venues including restaurants, night clubs and bars have been reported as common exposures in large outbreaks and clusters of COVID-19 (Furuse et al., 2020; ECDC, 2020; Lu et al., 2020, Fisher et al., 2020). Outbreaks have also been reported in some occupational settings including factories and warehouses (ECDC, 2020) and in educational settings (Ismail et al., 2020). However, information on the role any community setting has in facilitating transmission is still developing and limited (Lee et al., 2020).

The evidence on the risk of transmission in specific settings and activities is sparse but this information is crucial for evidence-based control measures for COVID-19. We investigated the association between occupation and infection among cases identified through the NHS Test and Trace programme in England through three retrospective case-control studies conducted monthly at the end of August, September and October 2020.

## Methods

### Study design and setting

We conducted three retrospective case-control studies, which were frequency-matched for age and geographical region. The study periods for the three studies were late August, late September, and late October 2020. The study population consisted of adults over 18 years old resident in England. Cases were people who had tested positive for SARS-CoV-2 and completed the National Health Service Test and Trace (NHS T&T) contact tracing questionnaire.

The controls were members of the public, registered as volunteers for a Market Research Panel, and who were not household contacts of a confirmed case. Controls were excluded if they had tested positive for SARS-CoV-2, or reported COVID-19 symptoms in the seven days prior to completing the survey. Figure 1 outlines the recruitment process and study flow.

**Figure 1.**
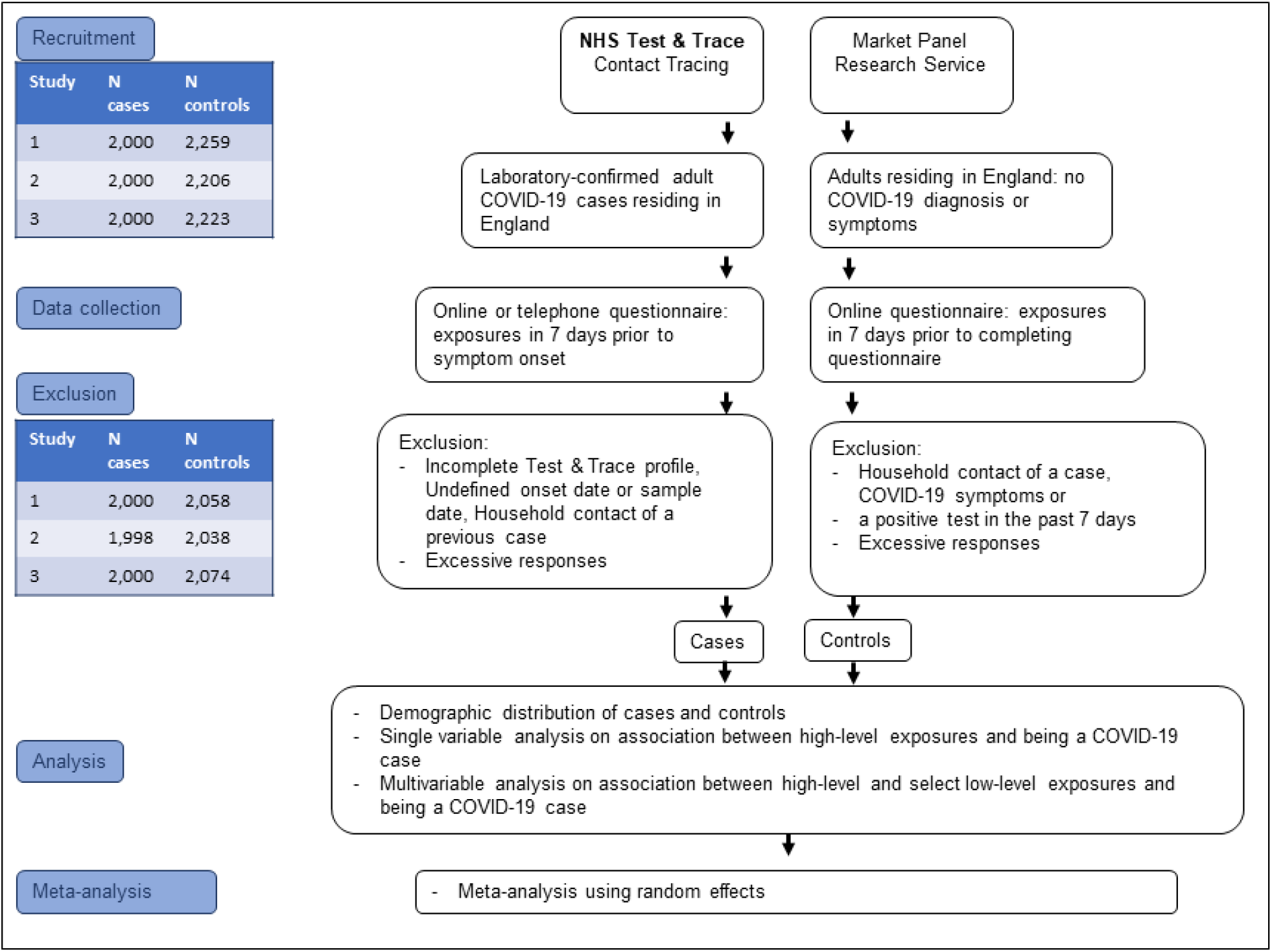
Study flow.

### Data collection

The data for the cases was collected through NHS T&T, where cases provided the information either through a digital route (self-completed) or through being interviewed over the phone. The NHS T&T assigns three different exposure levels for any given activity; the high-level categories being household or overnight stay, or work or leisure activity, with the following levels providing further granularity on the exposure reported. Controls completed an online survey with same exposure questions. However, they were not asked to provide details about their contacts for each exposure.

### Statistical analysis

Crude odds ratios (cORs) were obtained for each main exposure. Adjusted odds ratios (aORs) were obtained through multivariable analyses using penalised regression methods (see Firth, 1993). The main exposures for which the adjusted odds ratio showed evidence of an association were replaced in the multivariable analysis by their granular level exposures. Finally, we conducted a random-effects meta-analysis across the three studies to investigate how the association between exposure settings and the odds of infection differed in the three studies and to obtain pooled odds ratios (pORs).

## Results

### Descriptive analysis

Across the three studies, a total of 12,338 individuals were recruited, of which 6,000 were cases. Females were overrepresented with 6,926 (56%) participants, of which 3,268 (47%) were cases. There was a greater proportion of individuals in the control group that were of white ethnicity (83%) compared to the cases (65%), although ethnicity was not recorded in 9% of case respondents. A greater proportion of cases lived in areas of low deprivation (17%) than controls (12%), although deprivation score was unknown for a large proportion of control respondents (11%). Distribution was largely similar for all other demographic variables. (Table 1).

**Table 1.** Demographic distribution of cases and controls by study period.

|  | Period 1 |  |  |  | Period 2 |  |  |  | Period 3 |  |  |  |
| --- | --- | --- | --- | --- | --- | --- | --- | --- | --- | --- | --- | --- |
|  | Cases (n=2000) |  | Controls (n = 2058) |  | Cases (n = 2000) |  | Controls (n = 2206) |  | Cases (n = 2000) |  | Controls (n = 2074) |  |
|  | n | % | n | % | n | % | n | % | n | % | n | % |
| **Sex** |  |  |  |  |  |  |  |  |  |  |  |  |
| Male | 870 | 44% | 836 | 49% | 831 | 42% | 942 | 43% | 891 | 45% | 889 | 43% |
| Female | 1,095 | 55% | 1214 | 51% | 1104 | 55% | 1262 | 57% | 1069 | 53% | 1182 | 57% |
| Other sex | 0 | 0 | 8 | 0 | NA | NA | 2 | 0 | NA | NA | 3 | 0 |
| Missing sex | 35 | 2% | 0 | 0 | 65 | 3 | 0 | 0 | 40 | 2% | 0 | 0 |
| **Age group** |  |  |  |  |  |  |  |  |  |  |  |  |
| 18-27 | 697 | 35% | 694 | 34% | 658 | 33% | 730 | 33% | 490 | 24% | 505 | 24% |
| 28-37 | 478 | 24% | 536 | 26% | 451 | 23% | 493 | 22% | 458 | 23% | 471 | 23% |
| 38-47 | 338 | 17% | 349 | 17% | 309 | 15% | 420 | 19% | 368 | 18% | 411 | 20% |
| 48-57 | 294 | 15% | 256 | 12% | 344 | 17% | 237 | 11% | 408 | 20% | 315 | 15% |
| 58+ | 193 | 10% | 223 | 11% | 238 | 12% | 326 | 15% | 276 | 14% | 372 | 18% |
| Missing age | 0 | 0 | 0 | 0 | 0 | 0 | 0 | 0 | 0 | 0 | 0 | 0 |
| **Ethnicity** |  |  |  |  |  |  |  |  |  |  |  |  |
| White | 1,180 | 59% | 1625 | 79% | 1264 | 63% | 1839 | 83% | 1426 | 71% | 1804 | 87% |
| Mixed | 75 | 4% | 72 | 3% | 54 | 3% | 70 | 3% | 50 | 2% | 60 | 3% |
| Asian | 274 | 14% | 242 | 12% | 241 | 12% | 183 | 8% | 168 | 8% | 126 | 6% |
| Black | 78 | 4% | 87 | 4% | 79 | 4% | 87 | 4% | 32 | 2% | 63 | 3% |
| Other | 223 | 11% | 32 | 2% | 170 | 8% | 27 | 1% | 132 | 7% | 21 | 1% |
| Missing ethnicity | 170 | 8% | 0 | 0 | 192 | 10% | 0 | 0 | 192 | 10% | 0 | 0 |
| **Deprivation score** |  |  |  |  |  |  |  |  |  |  |  |  |
| 1 (most deprived) | 412 | 21% | 457 | 22% | 401 | 20% | 455 | 21% | 510 | 26% | 557 | 27% |
| 2 | 441 | 22% | 462 | 22% | 486 | 24% | 479 | 22% | 410 | 20% | 420 | 20% |
| 3 | 401 | 20% | 322 | 16% | 409 | 20% | 396 | 18% | 377 | 19% | 326 | 16% |
| 4 | 416 | 21% | 315 | 15% | 362 | 18% | 334 | 15% | 380 | 19% | 302 | 15% |
| 5 (least deprived) | 330 | 16% | 254 | 12% | 342 | 17% | 289 | 13% | 323 | 16% | 248 | 12% |
| Missing IMD | 0 | 0 | 248 | 12% | 0 | 0 | 253 | 11% | 0 | 0 | 221 | 11% |
| **Region** |  |  |  |  |  |  |  |  |  |  |  |  |
| East of England | 174 | 9% | 133 | 6% | 109 | 5% | 138 | 6% | 119 | 6% | 40 | 2% |
| East Midlands | 97 | 5% | 210 | 10% | 190 | 10% | 261 | 12% | 226 | 11% | 233 | 11% |
| London | 514 | 26% | 518 | 25% | 500 | 25% | 551 | 25% | 183 | 9% | 210 | 10% |
| North East | 85 | 4% | 118 | 6% | 108 | 5% | 151 | 7% | 111 | 6% | 286 | 14% |
| North West | 269 | 13% | 235 | 11% | 258 | 13% | 187 | 8% | 507 | 25% | 740 | 36% |
| South East | 293 | 15% | 241 | 12% | 291 | 15% | 243 | 11% | 163 | 8% | 110 | 5% |
| South West | 201 | 10% | 140 | 7% | 209 | 10% | 170 | 8% | 137 | 7% | 62 | 3% |
| West Midlands | 228 | 11% | 299 | 15% | 201 | 10% | 291 | 13% | 226 | 11% | 293 | 14% |
| Yorkshire and Humber | 139 | 7% | 164 | 8% | 134 | 7% | 214 | 10% | 328 | 16% | 100 | 5% |
|  | **2,000** |  | **2058** |  | **2000** |  | **2206** |  | **2000** |  | **2074** |  |

### Single variable analysis by study period

Working in social or home care, health care and working in hospitality were associated with being a case in all three study periods. Additionally, studies 2 and 3 showed evidence that working in a warehouse, emergency services, and close contact services were each associated with being a case. Furthermore, in these two periods, working or attending education was associated with increased odds of illness (Table 2).

**Table 2.** Single variable analysis results on occupational exposures by study period.

|  | Cases |  | Controls |  | Period 1 |  |  | Cases |  | Controls |  |  | Period 2 |  |  | Cases |  | Controls |  |  | Period 3 |  |  |
| --- | --- | --- | --- | --- | --- | --- | --- | --- | --- | --- | --- | --- | --- | --- | --- | --- | --- | --- | --- | --- | --- | --- | --- |
|  | Exposed<br>n | % | Exposed<br>n | % | Odds<br>ratio | 95%CI | P-value | Exposed<br>n | % | Exposed<br>n | % |  | Odds<br>ratio | 95%CI | P-value | Exposed<br>n | % | Exposed<br>n | % |  | Odds<br>ratio | 95%CI | P-value |
| Social care<sup>a</sup> | 169 | 8.5 | 39 | 1.9 | 4.78 | [3.33-6.99] | <0.001 | 95 | 4.8 | 21 | 1.0 |  | 4.38 | [2.74-7.26] | <0.001 | 52 | 2.6 | 25 | 1.2 |  | 2.19 | [1.33-3.69] | 0.001 |
| Health care<sup>b</sup> | 199 | 10.0 | 103 | 5.0 | 2.10 | [1.63-2.71] | <0.001 | 176 | 8.8 | 73 | 3.6 |  | 2.21 | [1.69-2.91] | <0.001 | 197 | 9.9 | 75 | 3.6 |  | 2.91 | [2.20-3.88] | <0.001 |
| Hospitality<sup>c</sup> | 98 | 4.9 | 54 | 2.6 | 1.91 | [1.35-2.73] | <0.001 | 91 | 4.6 | 34 | 1.7 |  | 2.61 | [1.76-3.96] | <0.001 | 86 | 4.3 | 47 | 2.3 |  | 1.94 | [1.33-2.84] | <0.001 |
| Close contact services<sup>d</sup> | 33 | 1.7 | 24 | 1.2 | 1.42 | [0.81-2.52] | 0.19 | 22 | 1.1 | 8 | 0.4 |  | 2.51 | [1.11-6.21] | 0.016 | 38 | 1.9 | 8 | 0.4 |  | 5.00 | [2.29-12.44] | <0.001 |
| Construction<sup>e</sup> | 77 | 3.9 | 75 | 3.6 | 1.06 | [0.76-1.48] | 0.73 | 99 | 5.0 | 40 | 2.0 |  | 1.85 | [1.31-2.63] | <0.001 | 154 | 7.7 | 59 | 2.8 |  | 2.85 | [2.08-3.94] | <0.001 |
| Warehouse<sup>f</sup> | 15 | 0.8 | 18 | 0.9 | 0.86 | [0.40-1.81] | 0.659 | 29 | 1.5 | 6 | 0.3 |  | 4.99 | [2.03-14.74] | <0.001 | 53 | 2.7 | 4 | 0.2 |  | 14.09 | [5.17-53.67] | <0.001 |
| Food production and agriculture<sup>g</sup> | 17 | 0.9 | 24 | 1.2 | 0.73 | [0.37-1.41] | 0.314 | 31 | 1.6 | 8 | 0.4 |  | 2.13 | [1.11-4.25] | 0.015 | 17 | 0.9 | 17 | 0.8 |  | 1.04 | [0.50-2.17] | 0.915 |
| Retail<sup>h</sup> | 74 | 3.7 | 116 | 5.6 | 0.64 | [0.47-0.87] | 0.004 | 68 | 3.4 | 60 | 2.9 |  | 0.89 | [0.63-1.25] | 0.479 | 73 | 3.7 | 74 | 3.6 |  | 1.02 | [0.73-1.44] | 0.888 |
| Transport<sup>i</sup> | 37 | 1.9 | 64 | 3.1 | 0.59 | [0.38-0.90] | 0.01 | 42 | 2.1 | 37 | 1.8 |  | 0.86 | [0.56-1.32] | 0.464 | 59 | 3.0 | 34 | 1.6 |  | 1.82 | [1.17-2.88] | 0.005 |
| Emergency services<sup>j</sup> | 21 | 1.1 | 37 | 1.8 | 0.58 | [0.32-1.02] | 0.045 | 10 | 0.5 | 7 | 0.3 |  | 1.46 | [0.50-4.53] | 0.44 | 25 | 1.3 | 10 | 0.5 |  | 2.61 | [1.21-6.11] | 0.008 |
| Arts, entertainment and recreation<sup>k</sup> | 18 | 0.9 | 41 | 2.0 | 0.45 | [0.24-0.80] | 0.004 | 27 | 1.4 | 30 | 1.5 |  | 0.74 | [0.43-1.26] | 0.239 | 30 | 1.5 | 17 | 0.8 |  | 1.84 | [0.98-3.57] | 0.042 |
| Work related travel<sup>l</sup> | 15 | 0.8 | 62 | 3.0 | 0.24 | [0.13-0.43] | <0.001 | 21 | 1.1 | 20 | 1.0 |  | 0.79 | [0.42-1.46] | 0.424 | 13 | 0.7 | 36 | 1.7 |  | 0.37 | [0.18-0.72] | 0.001 |
| Education<sup>m</sup> | 35 | 1.8 | 172 | 8.4 | 0.20 | [0.13-0.28] | <0.001 | 238 | 11.9 | 129 | 6.3 |  | 1.38 | [1.12-1.70] | 0.002 | 290 | 14.5 | 198 | 9.6 |  | 1.61 | [1.32-1.96] | <0.001 |
| Immigration<sup>n</sup> | 1 | 0.1 | 9 | 0.4 | 0.11 | [0.00-0.82] | 0.013 | 1 | 0.1 | 3 | 0.2 |  | 0.34 | [0.01-4.24] | 0.327 | 0 | 0.0 | 1 | 0.1 |  | 0.00 | [0.00-] | 0.326 |
| Military<sup>o</sup> | 0 | 0.0 | 11 | 0.5 | 0.00 | [0.00-0.36] | 0.001 | 11 | 0.6 | 3 | 0.2 |  | 3.76 | [0.99-21.01] | 0.029 | 10 | 0.5 | 3 | 0.1 |  | 3.47 | [0.89-19.64] | 0.044 |

- Working in social or domiciliary care – including care homes - Working in healthcare – including hospitals, GPs, drop-in clinics and other healthcare settings - Working in hospitality – including restaurants, food and drink outlets and lodging - Working in close contact services – barbers, hairdressers, nail salons, tattoo studios and tanning salons, and any other services which require close contact - Working in manufacturing or construction – including manufacturing of cars, electronics, textiles, pharmaceuticals, and construction labour or other construction professions - Working in warehouse settings – including working in warehouse, haulage, wholesale, or food distribution - Working in food production – including farming and agriculture, any food manufacturing and food production - Working in retail – working in retail stores and other retail related professions - Transport – Working in public transport including underground, trains, buses, and logistics and storage - Working in emergency services – including fire services, police and other emergency services - Working in arts, or recreation – including music, theatre, gyms, cinema, leisure centres - Work related travel – including attending conferences, door-to-door sales, visiting clients - Education – attending or working - Working in immigration or border force services – including office based and people facing professions - Working in the military – including the Navy, Army and Air Force

**Table 3.** Multivariable analysis results on occupational exposures by study period*.

| Work exposure | Period 1 |  |  |  | Period 2 |  |  |  | Period 3 |  |  |  |
| --- | --- | --- | --- | --- | --- | --- | --- | --- | --- | --- | --- | --- |
|  | Odds ratio | 95% C.I. |  | P-value | Odds ratio | 95% C.I. |  | P-value | Odds ratio | 95% C.I. |  | P-value |
| Warehouse<sup>a</sup> | 1.18 | 0.39 | 3.55 | 0.765 | 5.29 | 1.87 | 14.97 | 0.002 | 15.17 | 4.57 | 50.37 | <0.001 |
| Social care<sup>b</sup> | 5.68 | 3.52 | 9.18 | <0.001 | 4.94 | 2.85 | 8.56 | <0.001 | 2.42 | 1.34 | 4.35 | 0.003 |
| Hospitality<sup>c</sup> | 2.87 | 1.73 | 4.75 | <0.001 | 2.93 | 1.81 | 4.74 | <0.001 | 2.14 | 1.34 | 3.42 | 0.001 |
| Healthcare<sup>d</sup> | 2.83 | 1.98 | 4.05 | <0.001 | 2.73 | 1.94 | 3.83 | <0.001 | 3.09 | 2.20 | 4.35 | <0.001 |
| Construction<sup>e</sup> | 1.37 | 0.82 | 2.27 | 0.229 | 1.99 | 1.29 | 3.06 | 0.002 | 2.65 | 1.79 | 3.93 | <0.001 |
| Education<sup>f</sup> | 0.30 | 0.19 | 0.47 | <0.001 | 1.62 | 1.25 | 2.11 | <0.001 | 2.03 | 1.59 | 2.60 | <0.001 |
| Transport<sup>g</sup> | 1.07 | 0.57 | 1.99 | 0.838 | 0.78 | 0.47 | 1.30 | 0.348 | 2.13 | 0.91 | 5.00 | 0.081 |
| Emergency services<sup>h</sup> | 0.59 | 0.28 | 1.23 | 0.157 | 1.84 | 0.59 | 5.73 | 0.293 | 1.98 | 0.71 | 5.52 | 0.194 |
| Close contact services<sup>i</sup> | 2.92 | 1.11 | 7.68 | 0.03 | 1.11 | 0.40 | 3.04 | 0.843 | 1.24 | 0.83 | 1.86 | 0.293 |
| Retail<sup>j</sup> | 0.84 | 0.56 | 1.25 | 0.391 | 1.01 | 0.68 | 1.52 | 0.945 | 1.78 | 1.05 | 3.03 | 0.032 |
| Work - related travel<sup>k</sup> | 0.39 | 0.18 | 0.82 | 0.013 | 0.78 | 0.37 | 1.62 | 0.498 | 0.48 | 0.23 | 1.00 | 0.05 |
| Arts, entertainment, recreation<sup>l</sup> | 0.86 | 0.39 | 1.93 | 0.719 | 0.75 | 0.40 | 1.43 | 0.387 | 1.61 | 0.77 | 3.38 | 0.204 |
| Immigration services<sup>m</sup> | 1.34 | 0.07 | 25.60 | 0.845 | 0.30 | 0.03 | 3.52 | 0.337 | 2.60 | 0.10 | 66.58 | 0.564 |
| Military<sup>n</sup> | 0.05 | 0.00 | 3.45 | 0.162 | 6.53 | 1.16 | 36.81 | 0.033 | 4.80 | 0.86 | 26.89 | 0.075 |
| Food production and agriculture<sup>o</sup> | 1.20 | 0.41 | 3.54 | 0.742 | 1.84 | 0.85 | 3.97 | 0.121 | 0.90 | 0.36 | 2.24 | 0.814 |

\*Adjusted for age, sex, region, ethnicity, non-work exposures, and index of multiple deprivation, and leisure activities

### Multivariable analysis by study period

All multivariable analyses were adjusted for age, sex, ethnicity, socioeconomic deprivation (using index for multiple deprivation (IMD)), geographical region, and non-work community and leisure activities. All three studies provided strong statistical evidence of an association between working in healthcare, social care, or hospitality and becoming a COVID-19 case. Working in a warehouse, education or in construction were associated with increased odds of infection in the second and third study periods.

We also analysed leisure activities for both cases and controls. However, the results from these analyses were less consistent, apart from strong evidence that cases were more likely to engage in entertainment activities than controls (aORs range 5.38-11.64). In particular, going to pubs and bars was associated with increased odds of illness in the first and second study periods (aORs range 1.85-2.38). The full results from these analyses are provided in the supplementary evidence (SE 1).

The odds ratios for work exposures used in the meta-analyses were adjusted for age, sex, ethnicity, index for multiple deprivation, geographical region, and non-work community activities. There remained strong statistical evidence that working in healthcare (pOR 2.87, aOR range 2.72-3.08), social care (pOR 4.15, aOR range 2.46-5.41) or hospitality (pOR 2.36, aOR range 2.01-2.63) were associated with increased odds of being a COVID-19 case. There was evidence that working in warehouse setting was associated with increased odds (pOR 3.86, aOR range 1.06-14.19), however; there was a high heterogeneity between the three study periods (I^2^ = 78%, P=0.01), with a substantial increase in odds observed over the study periods. A similar pattern was also observed in education and construction. Figure 2 presents the full results.

**Figure 2.**
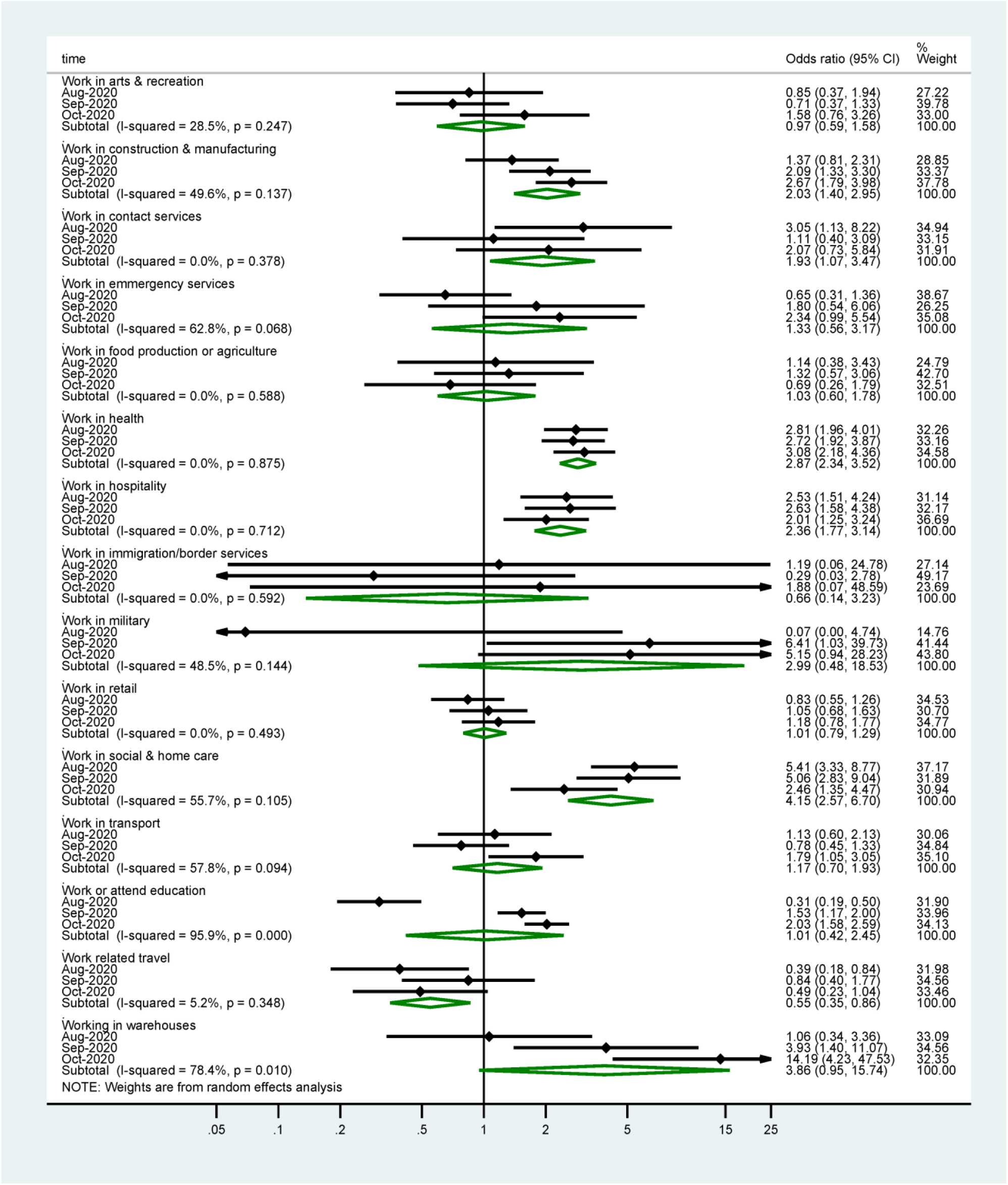
Meta-analysis of the three case-control study periods - grouped occupational exposures.

### Meta-analysis of the three case-control study periods – ungrouped occupational exposures

We explored the association between occupational exposures and infection using more granular information on work activities. Figure 3 presents the findings from these models. Cases had higher odds of working in warehouse than controls (pOR 5.55, aOR range 1.72-24.06). There was evidence that secondary school settings were associated with increased odds of being a COVID-19 case (pOR 2.56, aOR range 1.52-2.93). There was also strong statistical evidence across all studies that working in bars, restaurants and pubs was associated with elevated odds of being a case (pOR 2.86, aOR range 2.41-3.52). Working in hospital was linked with increased odds of COVID-19 (pOR 3.19, aOR range 2.29-4.06). There was also evidence of working in general practice surgeries (GPs) being associated with elevated odds of being a COVID-19 case (pOR 1.47, aOR range 0.51-2.71). However, there was substantial heterogeneity between studies for this setting (I^2^ 69.4%), with an increase in the adjusted odds ratios over the study periods.

**Figure 3.**
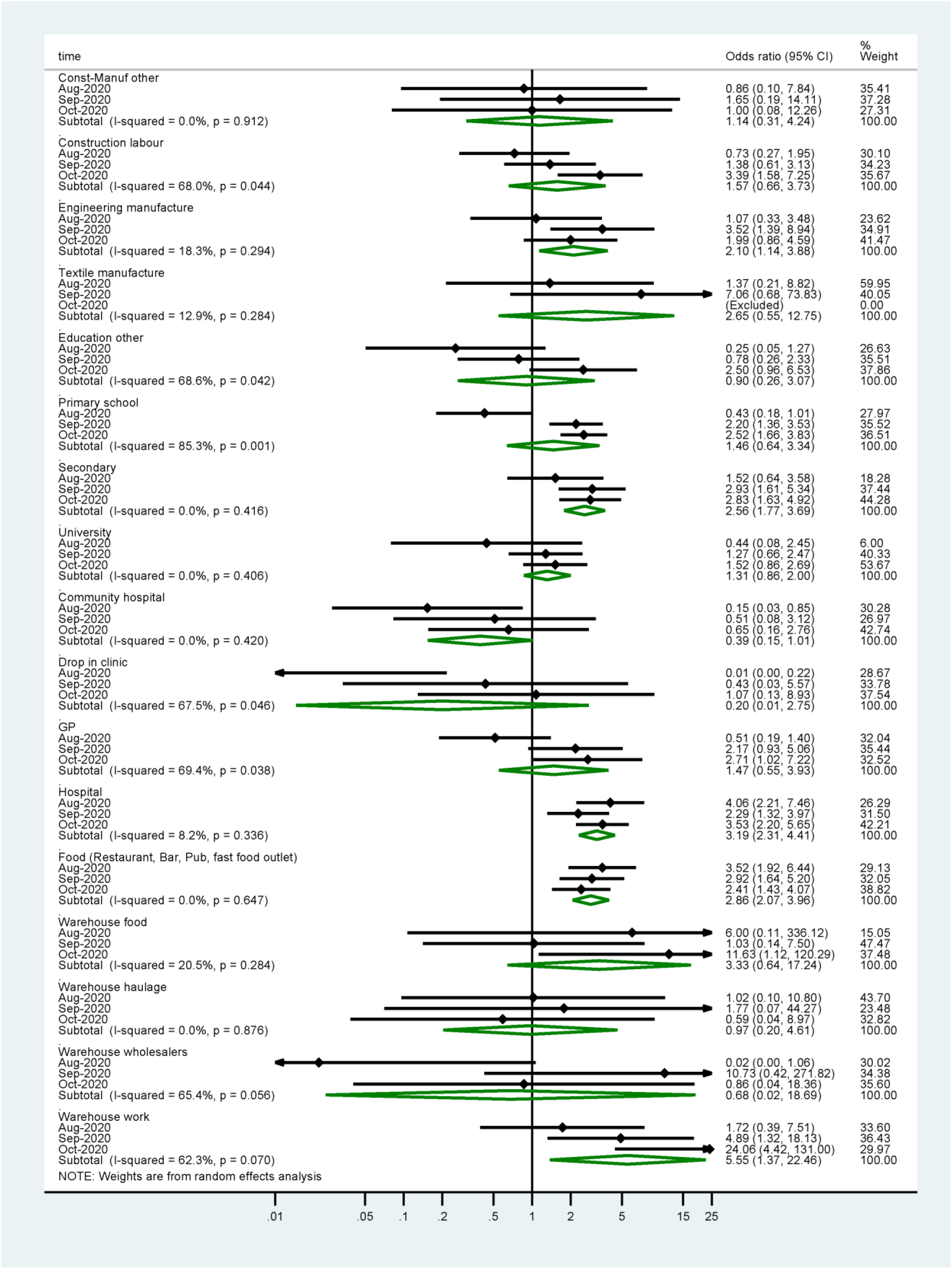
Meta-analysis of the three case-control study periods - ungrouped occupational exposures.

## Discussion

We observed strong associations between certain occupations and SARS-CoV-2 infection. The association with working in health and social care or in hospitality persisted across the three studies. Working in warehouse or in manufacturing and construction, or exposure to educational setting were associated with increased odds of being a COVID-19 case.

Our findings on the associations between infection and working in manufacturing, construction, and warehouse settings are consistent with the current evidence on most commonly reported outbreak settings (ECDC, 2020). The findings on educational settings are consistent with the recent evidence available on COVID-19 in these settings (Ismail et al., 2020). The findings regarding the association observed between working in health and social care settings and being a COVID-19 case were anticipated as these settings were closely associated with transmission in the first wave of the epidemic and, despite the appropriate use of personal protective equipment, healthcare workers experience high COVID-19 infection rates (Nguyen et al., 2020; Leclerc et al., 2020; ECDC, 2020). However, the findings from our studies highlight the risk associated with these settings, including hospitals and primary care settings like GPs.

Public health guidance and advice to reduce the risk of transmission in workplace settings have been published and disseminated (BEIS, 2020). It is likely that within the same occupational sector there will be examples of good and poor practice in the implementation of measures to mitigate transmission, and the risk of transmission is likely to vary from setting to setting within a sector. In addition, our study does not determine where each case was infected, or where transmission occurred, however, the associations we have observed are likely to reflect that it is more challenging to mitigate risk in some sectors compared to others. The observations presented here may be confounded by other factors including living arrangements, transport to work or socio-economic factors not explained by small area deprivation. For example, most of the cases who reported working in warehouses, or construction and manufacturing also resided in areas of high deprivation (results not shown) and it is possible that they may be more likely to live in multiple occupancy households, or travel together to work. However, the findings from this study provide useful evidence that can inform risk assessments and target public health action, especially as they provide data from a three-month period, with some consistent estimates of an association over time.

To the best of our knowledge, these are the only COVID-19 case-control studies conducted in England. As such, they provide a valuable insight into community risk factors for COVID-19 in England, and findings which are likely to be transferrable to other similar settings. They have provided timely evidence and allowed concurrent data collection from both cases and controls.

## Limitations

Limitations of the present study can be grouped into categories relating to means of data collection, sampling and selection bias, misclassification bias and confounding. Across all three studies, cases reported a smaller overall number of exposures than controls. The information from cases was collected at the point of contact tracing. It is possible that some cases may not have provided full or accurate information. One possible explanation for this is that the NHS T&T requires cases to record their contacts for each exposure, which substantially increases the time and effort required to record every exposure. This differential misclassification of exposure most likely biased the effect measures towards the null, hence the results described here are more likely to be underestimates than overestimates.

There is also potential for selection bias. Cases were randomly sampled from cases over 18 years of age in the NHS T&T system, which contains exposure information for all COVID-19 cases in England. Since approximately 85% of the cases transferred to the NHS T&T system are reached by the contact tracing programme (DHSC, 2020), there is likely to be some selection bias among cases if those who do not engage in contact tracing differ from the rest in terms of their exposures. Furthermore, the study does not include infected people who are not tested. The controls were sampled from a pool of volunteers using Market Research Panels, which most likely introduced some selection bias for controls. People registered on Market Research Panels may differ from the general population in their likelihood to engage in activities outside household setting. Furthermore, people registered on the Market Research Panel who chose to participate in the present study might differ from those registered on the Panel but who did not engage with the study. It is therefore likely that our sample of controls does not accurately represent the adult population of England. In addition, the present analyses could only control for confounding by exposures that data were available on, and therefore, residual confounding is likely to persist. For example, our analyses did not account for any behavioural change due to local restrictions.

## Conclusions

The studies identified that some workplace settings are associated with increased odds of being a case. However, it is not possible to determine how much of the transmission of SARS-CoV-2 took place within the workplace, and how much was associated with social, household or transport exposures so further work is needed to understand the risk of transmission in these setting.

## Data Availability

Applications for relevant anonymised data should be submitted to the Public Health England Office for Data Release: https://www.gov.uk/government/publications/accessing-public-health-england-data/about-the-phe-odr-and-accessing-data

