## Supplemental Table 1 for "Occupational exposures associated with being a COVID-19 case; evidence from three case-controls studies"

### SE-1: Table1: Multivariable analysis results on leisure activity exposures by study period

| | Period 1 | | | | Period 2 | | | | Period 3 | | | |
|---|---|---|---|---|---|---|---|---|---|---|---|---|
| **Leisure exposure** | Odds ratio | 95% C.I. | | P-value | Odds ratio | 95% C.I. | | P-value | Odds ratio | 95% C.I. | | P-value |
| Eating out[a] | 0.76 | 0.62 | 0.92 | 0.005 | 0.83 | 0.68 | 1.02 | 0.073 | 0.76 | 0.61 | 0.94 | 0.011 |
| Sports events[b] | 0.98 | 0.50 | 1.92 | 0.958 | 0.77 | 0.41 | 1.44 | 0.409 | 0.68 | 0.32 | 1.46 | 0.325 |
| Private events[c] | 1.40 | 0.80 | 2.44 | 0.233 | 1.72 | 0.86 | 3.42 | 0.122 | 0.73 | 0.24 | 2.21 | 0.578 |
| Attending places of worship[d] | 0.47 | 0.21 | 1.03 | 0.059 | 0.15 | 0.06 | 0.39 | 0 | 0.21 | 0.09 | 0.46 | 0 |
| Attending mass gatherings[e] | 0.93 | 0.36 | 2.40 | 0.881 | 2.45 | 0.60 | 10.02 | 0.211 | 18.74 | 0.75 | 467.26 | 0.074 |
| Charity events[f] | 0.50 | 0.20 | 1.25 | 0.14 | 0.29 | 0.11 | 0.77 | 0.013 | 0.40 | 0.13 | 1.24 | 0.113 |
| Personal exercise[g] | 0.66 | 0.49 | 0.89 | 0.006 | 0.96 | 0.72 | 1.27 | 0.773 | 0.88 | 0.67 | 1.15 | 0.347 |
| Personal care[h] | 0.16 | 0.09 | 0.30 | <0.001 | 0.41 | 0.25 | 0.69 | 0.001 | 0.43 | 0.26 | 0.71 | 0.001 |
| Shopping[i] | 0.13 | 0.11 | 0.16 | <0.001 | 0.15 | 0.13 | 0.17 | 0 | 0.17 | 0.14 | 0.20 | 0 |
| Travel[j] | 1.01 | 0.67 | 1.53 | 0.956 | 0.76 | 0.48 | 1.20 | 0.241 | 0.42 | 0.24 | 0.72 | 0.002 |
| Visiting family and friends[k] | 0.27 | 0.21 | 0.34 | <0.001 | 0.34 | 0.27 | 0.43 | 0 | 0.38 | 0.29 | 0.50 | 0 |
| Healthacare (non-Covid19 reason)[l] | 0.23 | 0.14 | 0.37 | <0.001 | 0.30 | 0.20 | 0.44 | 0 | 0.27 | 0.18 | 0.39 | 0 |
| Entertainment - other[m] | 11.64 | 4.03 | 33.65 | <0.001 | 5.38 | 1.95 | 14.85 | 0.001 | 9.41 | 2.19 | 40.45 | 0.003 |
| Entertainment - pubs and bars[n] | 1.85 | 1.10 | 3.11 | 0.02 | 2.38 | 1.34 | 4.23 | 0.003 | 1.86 | 0.77 | 4.52 | 0.171 |

a. *Eating out – in different restaurants, cafes, pubs*
b. *Sports events - including football, cricket, rugby, tennis etc. matches, horse races and other sports events*
c. *Private events – including weddings, funerals, parties, other social gatherings*
d. *Places of worship – including churches, mosques, synagogues and other places of worship*
e. *Mass gatherings – raves, gigs, festivals, protests and live music events*
f. *Charity and community – fundraising, volunteering, bootsale, corporate events, educational classes*
g. *Personal exercise – e.g. at the gym, swimming, running clubs, casual exercise with friends*
h. *Personal care - Visiting nail salons, hairdressers, barbers, tanning and tattoo studios*
i. *Shopping - including shopping at supermarkets, local shops and pharmacies*
j. *Travel - any international and domestic travel including trains and flights*
k. *Visiting family or friends – indoors or outdoors*
l. *Health care - visiting healthcare services for non-COVID-19 reasons*
m. *Other entertainment activities (non-specific)*
n. *Going to pub or bar (entertainment, not e.g. eating out)*
